# Association of Lung Fibrotic Changes and Cardiological Dysfunction with Hypertension in Long COVID-19 cohort

**DOI:** 10.1101/2022.06.13.22276244

**Authors:** Ainur Tauekelova, Zhanar Kalila, Bakhtiyar Akerke, Zarina Sautbayeva, Polina Len, Aliya Sailybayeva, Sadyk Khamitov, Nazira Kadroldinova, Natalie S. Barteneva, Makhabbat Bekbossynova

**Affiliations:** National Research Center for Cardiac Surgery, Nur-Sultan, Kazakhstan; School of Sciences and Humanities, Nazarbayev University, Nur-Sultan, Kazakhstan; School of Medicine, Nazarbayev University, Nur-Sultan; Harvard Medical School, Brigham and Women’s Hospital, Boston, MA, United States

**Keywords:** LongCOVID-19, post-COVID-19, hypertension, diabetes, ferritin, fibrinogen, lung sequelae, cardiovascular sequalae, female gender

## Abstract

**Background:** Long COVID-19 symptoms appeared in many COVID-19 survivors. However, the prevalence and symptoms associated with long COVID and its comorbidities have not been established.

**Methods:** Between May and September 2020, we included 312 patients with post-COVID-19 from 21 primary care centers if they had any persistent symptoms for at least three months from the first onset of the disease. On the 6 months follow up, their lung function was assessed by CT and spirometry, whereas cardiac function was assessed by electrocardiogram (ECG), Holter ECG, Echocardiography, and 24-hour blood pressure monitoring. A six-minute test (6MWT) was conducted on 308 participants during the follow-up visit. All participants were given a questionnaire with items on demographic information, current complaints, comorbidities, and medications, and Chalder Fatigue Scale (CFS) questionnaire. Statistical analysis was done using R vs. 4.1.2. Two-group comparison of continuous variables was performed using a T-test for normally distributed data, and the Mann-Whitney Wilcoxon test, ANOVA, and Kruskal-Wallis tests were applied for multiple comparisons following with Tukey and Dunn tests as post-hoc methods. Hochberg p-value adjustment was used to reduce the false discovery rate during multiple comparisons. Categorical variables were analyzed with Fisher’s Exact test.

**Results:** Of 312 persons investigated, there was no significant gender difference between post-COVID-19 clinical manifestations except for memory dysfunction and anxiety, more prevalent among female participants. Chalder Fatigue Score ≥4 was predominant in female participants (243, 78%). 39 (12.5%) participants reported having type 2 diabetes mellitus, and 158 (50.64%) had hypertension.

Among the tested parameters, those positively correlated with comorbid conditions include age, BMI, D-dimers, NT-proBNP, C-reactive protein, neutrophils, fasting glucose, and HbA1c; hypertension also shows three associations that were not found in patients when examining the role of diabetes: increased hemoglobin, fibrinogen, and ferritin. 24-hour blood pressure monitoring revealed significantly higher systolic and diastolic blood pressure, left ventricular hypertrophy, and elevated NT-proBNP in participants with hypertension and subjects with type 2 diabetes. Left ventricular diastolic dysfunction is more frequently present in patients with hypertension.

Chest CT was conducted on 227 (72.8%) participants 5.8±0.9 months after the onset of COVID-19. The most common registered CT abnormality was chronic bronchitis (198, 87.2%), followed by fibrotic changes in (83, 36.6%) and mediastinal lymphadenopathy (23, 10.1%).

Immunological test results showed that SARS-CoV19 IgG antibodies were present in 241 subjects (77.2%), and SARS-CoV19 IgM antibodies were present in 9 subjects (2.88%).

**Conclusions:** Our study provides valuable clues for long-term post-sequelae in a cohort of the Long COVID-19 subjects. We demonstrated a strong association of signs of cardiac dysfunction, lung fibrotic changes, increased hemoglobin, fibrinogen, and ferritin with hypertension but not with other comorbidities. Our results are of importance for understanding the Long Covid-19 syndrome.

## Introduction

The COVID-19 disease, resulting from infection with SARS-CoV-2, is highly contagious and may spread asymptomatically or presymptomatically (Furukawa et al., 2020; Rivett et al., 2020; Buitrago-Garci et al., 2020). There is a growing concern regarding the potential for COVID-19 infection to contribute to a burden of chronic cardiovascular and respiratory symptoms among recovered individuals (Leth et al., 2021). The SARS-CoV2 virus appears unique among coronaviruses due to the high rate of transmission and atypical pattern of inflammatory response leading in severe cases to cytokine storm phenomenon (rev. Tang et al., 2020). Long-COVID-19 or post-COVID-19 syndrome describes symptoms lasting for more than three months after the first COVID-19 symptoms onset (Yong, 2021), and there is a critical need to evaluate and understand potential long-term implications of COVID-19. The most frequent symptoms are shortness of breath and fatigue (Cares-Marambio et al., 2021; Shah et al., 2021; Davido et al., 2020; Halpin et al., 2020).

Many meta-analyses identified that the elderly population with pre-existing disease conditions such as type 2 diabetes mellitus (Singh et al., 2020; Kumar et al., 2020) and hypertension (Pranata et al., 2020; de Almeida-Pititto et al., 2021) are found to be at an increased risk of getting COVID-19 infection and its complications (Deng et al., 2020). COVID-19 has been linked to multiple extra-respiratory symptoms more prevalent in patients with positive cardiovascular disease history (Han et al., 2020). Long-lasting alternations in pulmonary function, including the reduced diffusing capacity of lungs and the restrictive syndrome, were observed in patients surviving COVID-19 pneumonia during a 6-month and 12 months follow-up period (Fortini et al., 2021; Fumagalli et al., 2021; Wu et al., 2021; Ordinola Navarro et al., 2021). However, long COVID affects even mild-to-moderate cases of survivors of COVID-19 with pulmonary sequelae reported after recovery (Wang et al., 2020).

In this paper, we report results from a prospective cohort study of 312 patients with COVID-19 onset between May and September 2020. We are aimed to determine the prevalence and extent of allowing analysis of persisting Long COVID-19 symptoms over 6 months duration emphasizing the patient’s cardiovascular and lung sequelae of COVID-19.

## 2. Materials and Methods

### 2.1 Study design and participants

This is a prospective cohort study that was based on a pilot project of the post-COVID center in JSC “National Research Cardiac Surgery Center” in Nur-Sultan, Kazakhstan. The study was conducted in December 2020. The post-COVID center was established according to the order of the Ministry of Health of the Republic of Kazakhstan No. 763 dated November 24, 2020, as a pilot project for the improvement of the coronavirus infection situation in the healthcare of the country. Participants were referred from the primary care centers of Nur-Sultan (Kazakhstan) to the post-COVID center. Participants were selected if they had any persistent symptoms for at least three months from the first onset of the disease. Selected 312 were referred to the follow-up visit to JSC “National Research Cardiac Surgery Center.” All participants had confirmed infection with positive PCR during the active infection or laboratory immunohistochemical results. The study was ethically approved according to the order of the Ministry of Health of the Republic of Kazakhstan, and informed consent was taken in the primary care centers.

### 2.2 Clinical Assessment

All participants were given a questionnaire with items on demographic information, current complaints, comorbidities, and medications (**Appendix 1**). All participants were given Chalder Fatigue Scale (CFS) questionnaire (**Appendix 2**) for completion. Vital signs of participants, including blood pressure and heart rate were performed by one of three physicians participating in data collection. Venous blood samples were collected from all participants for complete blood count, including hemoglobin, white blood cells, lymphocytes, neutrophils, and platelet count, for biochemistry panel, including serum creatinine, blood urea, total cholesterol, glucose, and ALT, for coagulation profile, including fibrinogen and D-Dimer (DD), and for glycohemoglobin A1c (HbA1c), C-reactive protein, ferritin, vitamin B12, vitamin 25(OH)D, and NT-proBNP. For coagulation tests AVATUBE R with sodium citrate was used, and Sysmex CS-2500 instrument (Sysmex, Japan). Blood for DD was collected in AVATUBE R with sodium citrate, and D-dimers quantification was performed with Cobas R 6000. Additionally, blood analysis for immunological tests was performed, including SARS-CoV-2 IgG, SARS-CoV-2 IgM, CMV IgG, and CMV IgM antibodies quantified by ELISA. All participants were proposed chest CT, but 85 participants (28.3%) refused due to personal claims. Cardiac function was assessed by electrocardiogram (ECG), Holter ECG, Echocardiography, and 24-hour blood pressure monitoring in all participants. A six-minute test (6MWT) was conducted in 308 participants at the follow-up visit. The medical records of each participant were reviewed independently by three physicians (cardiologist with more than 10 years of experience) of the hospital. All CT images were reviewed in random order by one senior cardiothoracic radiologist who were not aware of any clinical or laboratory findings or participant outcomes.The confirmation of COVID-19 infection was done by RT-PCR assay on nose/throat swab or sputum samples using CFX96 R Real-Time System (Bio-Rad Laboratories, Inc., USA).

### 2.3 Statistical Analysis

Statistical analysis was performed in parallel using R vs. 4.1.2 (2021-11-01) and GraphPad Prism vs. 8.4.3 (Dotmatics, USA). Normality was assessed using the Shapiro test. Two-group comparison of continuous variables was performed using T-test for normally distributed data and MannWhitney Wilcoxon test if the condition of normality was violated. For multiple comparisons ANOVA and Kruskal-Wallis tests were applied with Tukey and Dunn tests as post-hoc methods. Hochberg p-value adjustment was used to reduce the false discovery rate during multiple comparisons. Categorical variables were analyzed with Fisher’s Exact test.

### 2.5 Ethical considerations

All participants of the study provided written informed consent. The study was approved by the National Research Center Cardiac Surgery Center Ethical Committee (#01-97/2021 from 22/04/21). No compensation was provided to participants (only reimbursing of travelling costs).

## 3. Results

### 3.1 Cohort description

The comorbidities are reported in **Table 1** from the most prevalent to the least. Overall, 223 participants had at least one comorbidity. The most common comorbidities included hypertension (154, 50.6%), gastrointestinal (GI) disorders (74, 23.73%), and neurological disorders (43, 13.78%). Some of the participants also had overlapping of the comorbidities.

**Table 1.**
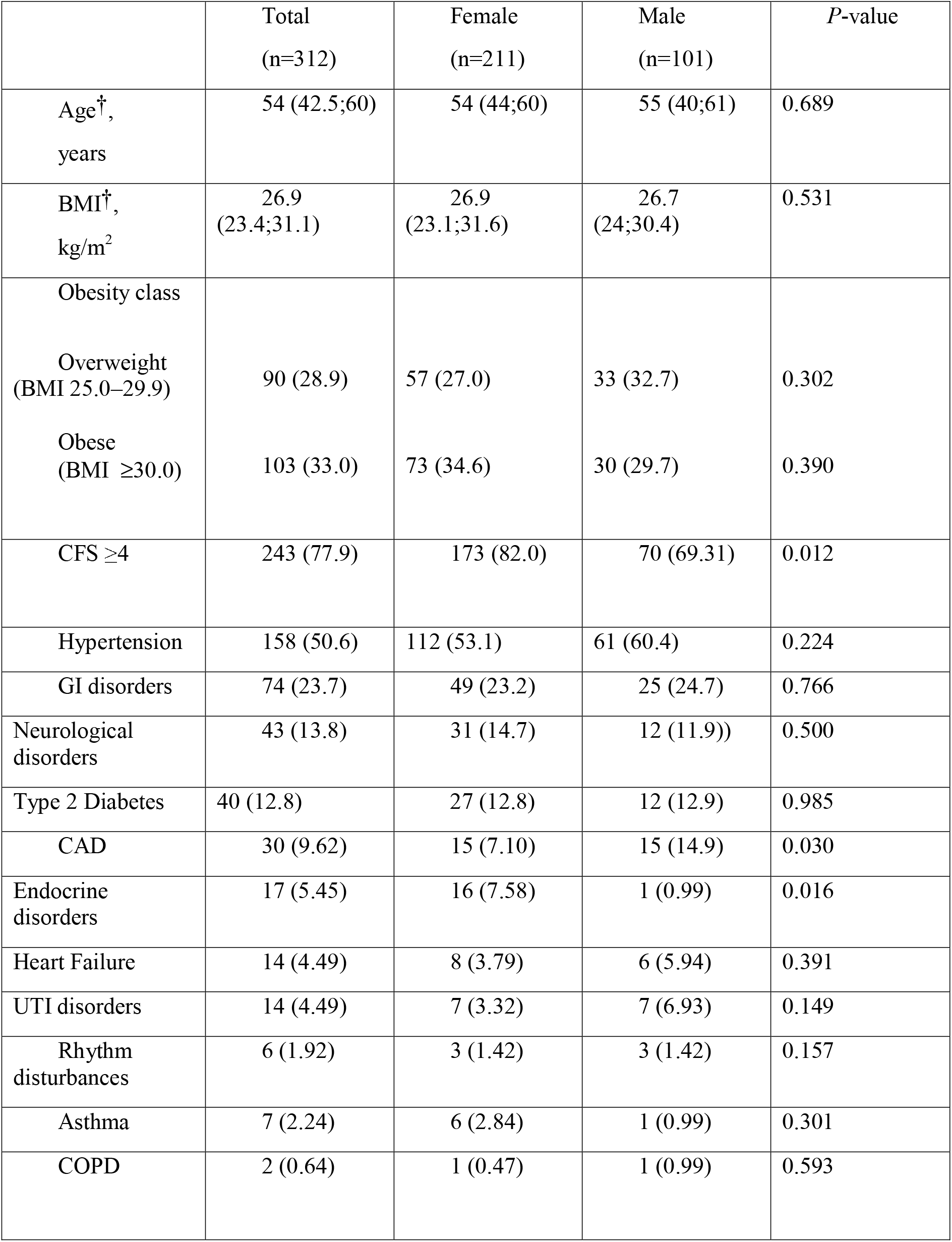
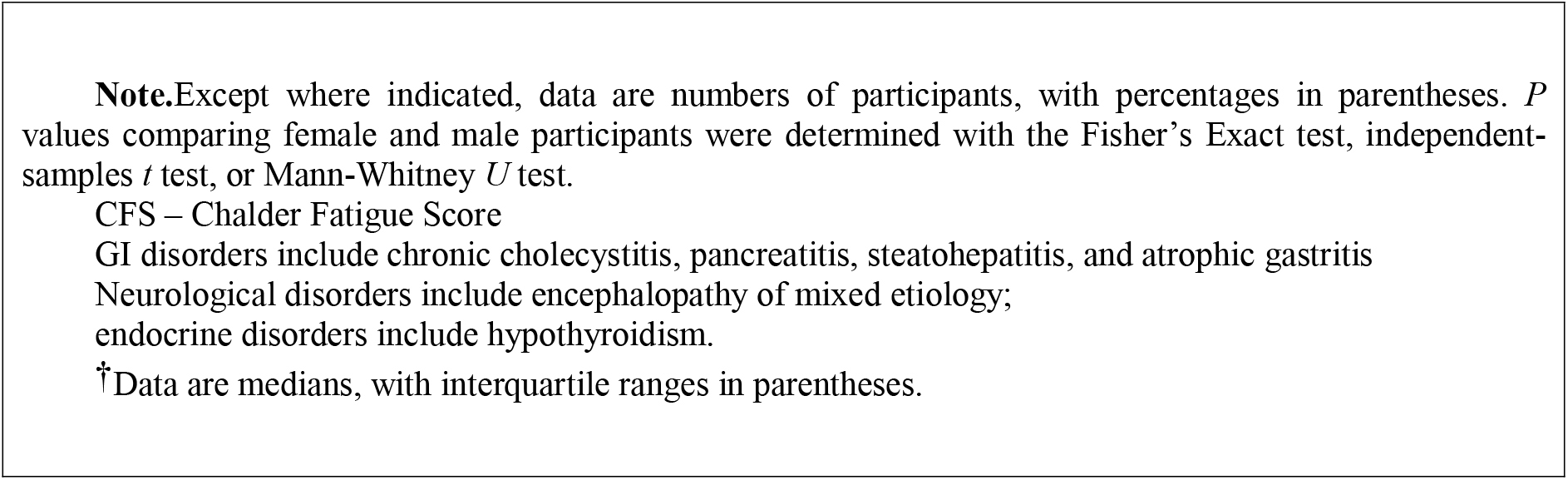
Clinical characteristic and comorbidities in participants after COVID-19 infection.

|  | Total<br>(n=312) | Female<br>(n=211) | Male<br>(n=101) | *P*-value |
| --- | --- | --- | --- | --- |
| Age<sup>†</sup>,<br>years | 54 (42.5;60) | 54 (44;60) | 55 (40;61) | 0.689 |
| BMI<sup>†</sup>,<br>kg/m<sup>2</sup> | 26.9<br>(23.4;31.1) | 26.9<br>(23.1;31.6) | 26.7<br>(24;30.4) | 0.531 |
| Obesity class |  |  |  |  |
| Overweight<br>(BMI 25.0–29.9) | 90 (28.9) | 57 (27.0) | 33 (32.7) | 0.302 |
| Obese<br>(BMI ≥30.0) | 103 (33.0) | 73 (34.6) | 30 (29.7) | 0.390 |
| CFS ≥4 | 243 (77.9) | 173 (82.0) | 70 (69.31) | 0.012 |
| Hypertension | 158 (50.6) | 112 (53.1) | 61 (60.4) | 0.224 |
| GI disorders | 74 (23.7) | 49 (23.2) | 25 (24.7) | 0.766 |
| Neurological<br>disorders | 43 (13.8) | 31 (14.7) | 12 (11.9)) | 0.500 |
| Type 2 Diabetes | 40 (12.8) | 27 (12.8) | 12 (12.9) | 0.985 |
| CAD | 30 (9.62) | 15 (7.10) | 15 (14.9) | 0.030 |
| Endocrine<br>disorders | 17 (5.45) | 16 (7.58) | 1 (0.99) | 0.016 |
| Heart Failure | 14 (4.49) | 8 (3.79) | 6 (5.94) | 0.391 |
| UTI disorders | 14 (4.49) | 7 (3.32) | 7 (6.93) | 0.149 |
| Rhythm<br>disturbances | 6 (1.92) | 3 (1.42) | 3 (1.42) | 0.157 |
| Asthma | 7 (2.24) | 6 (2.84) | 1 (0.99) | 0.301 |
| COPD | 2 (0.64) | 1 (0.47) | 1 (0.99) | 0.593 |

**Note.** Except where indicated, data are numbers of participants, with percentages in parentheses. *P* values comparing female and male participants were determined with the Fisher's Exact test, independent-samples *t* test, or Mann-Whitney *U* test.
CFS – Chalder Fatigue Score
GI disorders include chronic cholecystitis, pancreatitis, steatohepatitis, and atrophic gastritis
Neurological disorders include encephalopathy of mixed etiology;
endocrine disorders include hypothyroidism.
†Data are medians, with interquartile ranges in parentheses.

The proportion of coronary artery disease (CAD) in male participants was higher than in females, 14.85% in males vs. 7.11% in females, P=0.030. Meanwhile, endocrine disorders were more prevalent among female participants (16, 7.58%) vs. 1, 0.99%), respectively, P=0.016).

Respiratory comorbidities include asthma and chronic obstructive disease (COPD) (9, 2.88%) with no significant difference among female and male participants.

A total of 312 participants who were previously infected with COVID-19 infection were included in the study. Mean time for the post-COVID-19 assessment was 5.8 ±0.9 months onset of first symptoms of COVID-19 infection. 111 participants had hospitalization during the active COVID-19 infection while 201 participants were treated at home. Clinical characteristics of the participants are reported in **Table 1**.

Median age of all participants were 54 years (IQR, 17.5) and 211 participants (67.6%) were females. One participant required extracorporeal oxygenation membrane oxygenation (ECMO) during the active infection. Median age of all participants was 54 years (IQR, 17.5) and 211 participants (67.6%) were females. Median BMI of all participants was 26.9 kg/m^2^ (IQR, 7.7) with 90 participants (28.9%) overweight and 103 participants (33.0%) obese. Chalder Fatigue Score ≥4 was present in 243 participants (78%) with more predominance in female participants.

### 3.2 Clinical manifestations at the follow-up and laboratory results

Clinical manifestations of participants at the follow-up are reported in **Table 2**. The symptoms are presented from the most common to the least. The most common symptoms among 312 participants were fatigue (220, 70.51%), tiredness (180, 57.69%), and sleep disturbances (168, 53.85%), muscle pain (109, 34.9%), memory disfunction (108, 34.9%), dizziness (34.9%), headache (79, 25.3%), instability of blood pressure mostly with increased systolic blood pressure (62, 19.9%), palpitation (47, 15.1%), dyspnea on exertion (45, 14.4%), joint pain (39, 12.5%), and increased sweating (39, 12.5). Overall, there were no significant difference between post-COVID-19 clinical manifestations among female and male participants, except memory dysfunction and anxiety. Both were more prevalent among female participants, 81 female participants (38.4%) reported memory dysfunction (27 male participants (26.7%), *P*=0.043) and 28 female participants (12.3%) reported anxiety (2 male participants (1.98%), *P*=0.003).

**Table 2.** Participants-reported clinical manifestations persistent after COVID-19 infection at median of 6 months follow-up.

|  | Total<br>(n=312) | Female<br>(n=211) | Male<br>(n=101) | *P*-value |
| --- | --- | --- | --- | --- |
| Fatigue | 220 (70.5) | 143 (67.8) | 77 (76.2) | 0.125 |
| Tiredness | 180 (57.7) | 118 (55.9) | 62 (61.5) | 0.361 |
| sleep<br>disturbances | 168 (53.9) | 116 (55.0) | 52 (51.5) | 0.563 |
| Muscle<br>pain | 109 (34.9) | 78 (37.0) | 31 (30.7) | 0.277 |
| memory dysfunction | 108 (34.6) | 81 (38.4) | 27 (26.7) | 0.043 |
| dizziness | 107 (34.3) | 80 (37.9) | 27 (26.7) | 0.052 |
| headache | 79 (25.3) | 59 (28.0) | 20 (19.8) | 0.121 |
| BP instability | 62 (19.9) | 38 (18.0) | 24 (23.8) | 0.233 |
| Palpitation | 47 (15.1) | 34 (16.1) | 13 (12.9) | 0.454 |
| Dyspnea on exertion | 45 (14.4) | 31 (14.7) | 14 (13.9) | 0.845 |
| joint pain | 39 (12.5) | 30 (14.2) | 9 (8.9) | 0.185 |
| increased sweating | 39 (12.5) | 29 (13.7) | 10 (9.9) | 0.337 |
| anxiety | 28 (8.97) | 26 (12.32) | 2 (1.98) | 0.003 |
| hair loss | 27 (8.65) | 19 (9.00) | 8 (7.92) | 0.750 |
| Dyspnea on rest | 18 (5.77) | 11 (5.21) | 7 (6.93) | 0.543 |
| decreased vision | 18 (5.77) | 12 (5.69) | 6 (5.94) | 0.928 |
| GI symptoms | 16 (5.13) | 12 (5.69) | 4 (3.96) | 0.518 |
| chest pain in rest | 12 (3.85) | 11 (5.21) | 1 (0.99) | 0.070 |
| Daytime sleeping | 11 (3.53) | 6 (2.84) | 5 (4.95) | 0.563 |
| Cough with sputum | 9 (2.88) | 7 (3.32) | 2 (1.98) | 0.509 |
| mood change | 8 (2.56) | 5 (2.37) | 3 (2.97) | 0.753 |
| chest pain on exertion | 7 (2.24) | 5 (2.37) | 2 (1.98) | 0.828 |
| dry cough | 7 (2.24) | 7 (3.32) | 0 | 0.064 |
| numbness in extremity | 7 (2.24) | 5 (2.37) | 2 (1.98) | 0.828 |
| decreased hearing | 6 (1.92) | 6 (2.84) | 0 | 0.087 |
| tremor in extremity | 5 (1.60) | 3 (1.42) | 2 (1.98) | 0.713 |
| Impaired sense of smell | 5 (1.60) | 3 (1.42) | 2 (1.98) | 0.713 |
| increased weight | 4 (1.28) | 3 (1.42) | 1 (0.99) | 0.751 |
| decreased concentration | 3 (0.96) | 2 (0.95) | 1 (0.99) | 0.971 |
| Wheezing | 2 (0.67) | 1 (0.47) | 1 (0.99) | 0.593 |
| nervousness | 2 (0.64) | 2 (0.95) | 0 | 0.326 |
| presyncope | 2 (0.64) | 2 (0.95) | 0 | 0.326 |
| syncope | 2 (0.64) | 2 (0.95) | 0 | 0.593 |
| Impaired sense of taste | 2 (0.64) | 1 (0.47) | 1 (0.99) | 0.593 |
| fever | 2 (0.64) | 1 (0.47) | 1 (0.99) | 0.593 |
| edema of extremities | 2 (0.64) | 2 (0.95) | 0 | 0.326 |
| Note.— Data are numbers of participants, with percentages in parentheses. *P* values comparing female and male participants were determined with the $\chi^2$ test.<br>BP – blood pressure |  |  |  |  |

Out of 312 participants, 148 (47.44%) had neither hypertension nor type 2 diabetes mellitus, 33 (10.58%) had both hypertension and diabetes, 125 (40.06%) had hypertension only, and 6 (1.92%) participants had diabetes only. Overall, 39 (12.5%) participants reported having type 2 diabetes mellitus, and 158 (50.64%) had hypertension. Among the tested parameters, those positively correlated with comorbid conditions include age, BMI, D-dimers, NT-proBNP, C-reactive protein, neutrophils, fasting glucose, and HbA1c; hypertension also shows three associations that were not found in patients when examining the role of diabetes: increased hemoglobin, fibrinogen, and ferritin (**Figures 2-4**).

**Figure 1.**
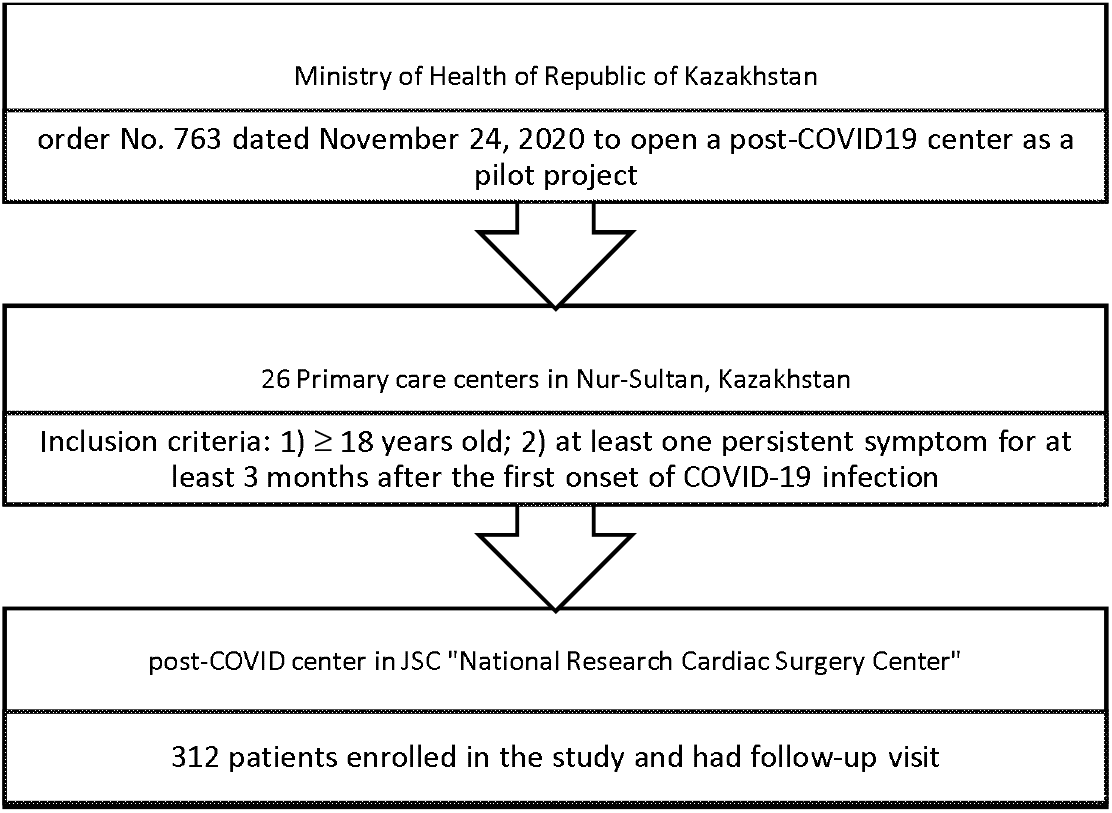
Study flow diagram.

**Figure 2.**
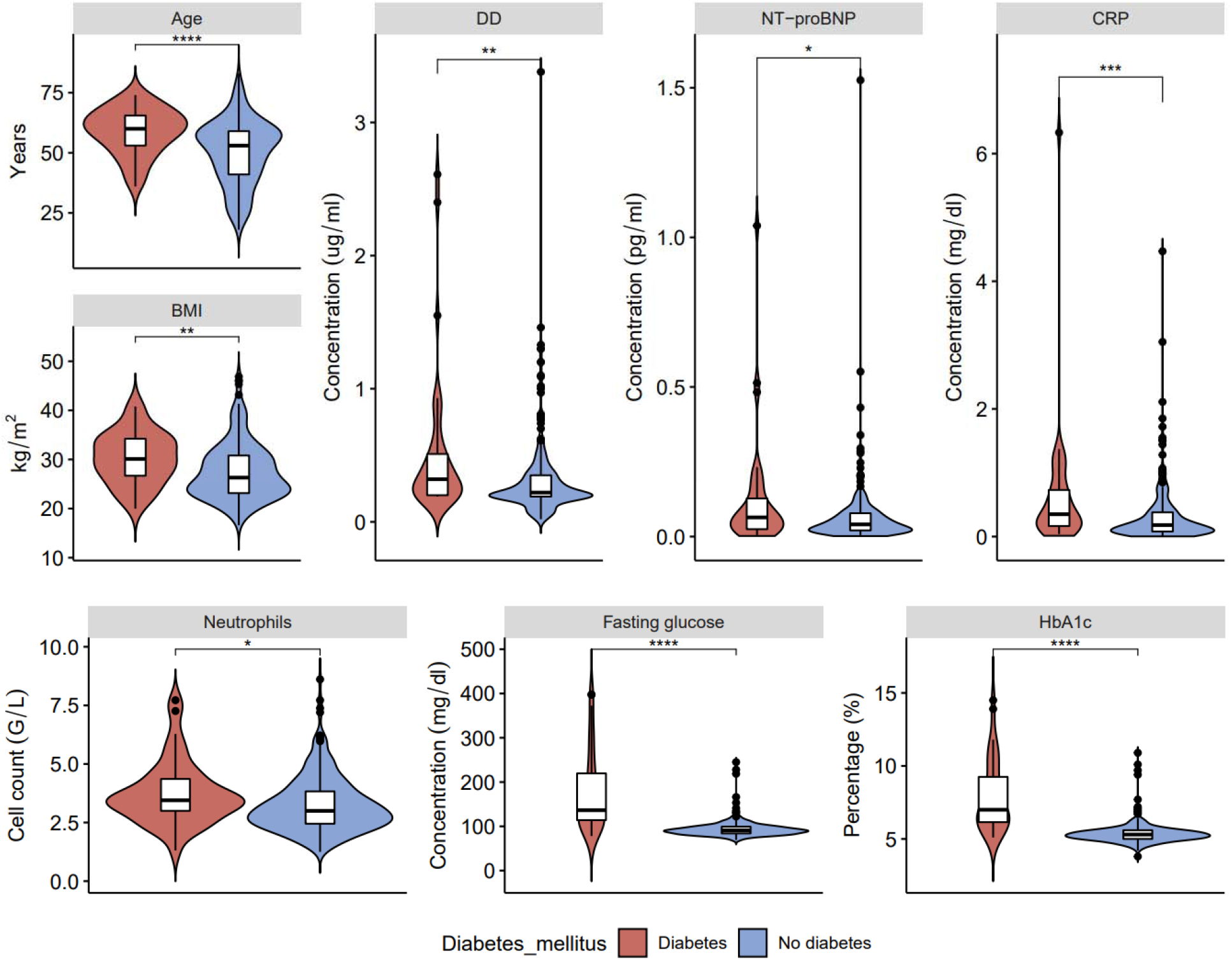
Clinical characteristics and laboratory results of blood samples in post-COVID-19 patients with and without type 2 diabetes: age, D-dimers (DD), ventricular natriuretic peptide (NT-proBNP), C-reactive protein (CRP), body mass index (BMI), neutrophils, fasting glucose and glycated hemoglobin (HbA1c) (\**p* < 0.05, \*\**p* < 0.01, \*\*\**p* < 0.001, \*\*\*\**p* < 0.0001).

**Figure 3.**
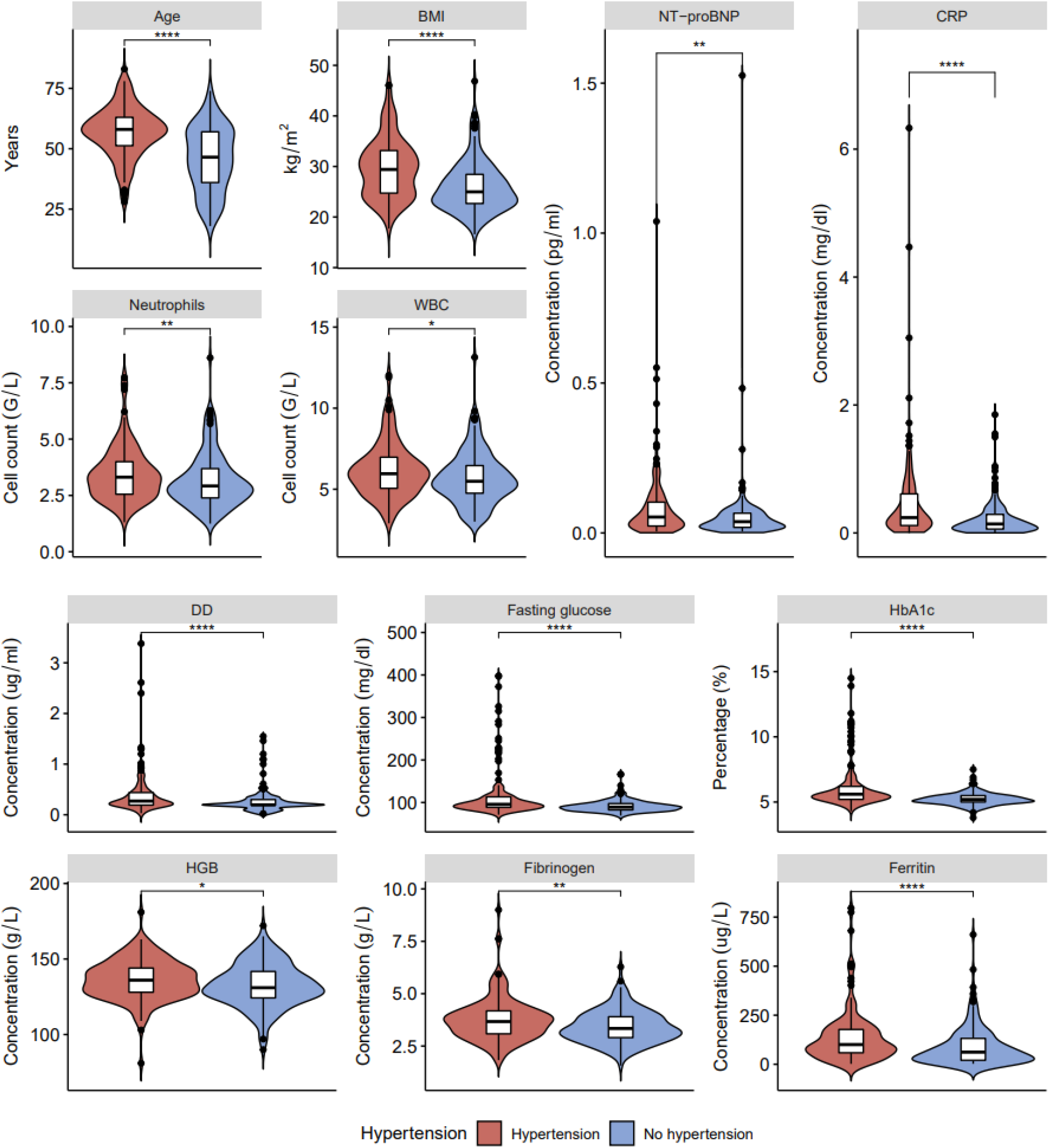
Clinical characteristics and laboratory results of blood samples in post-COVID-19 patients with and without hypertension: age, BMI, NT-proBNP, CRP, neutrophils, white blood cells (WBC), DD, fasting glucose, HbA1c, HGB, fibrinogen and ferritin (\**p* < 0.05, \*\**p* < 0.01, \*\*\*\**p* < 0.0001).

**Figure 4.**
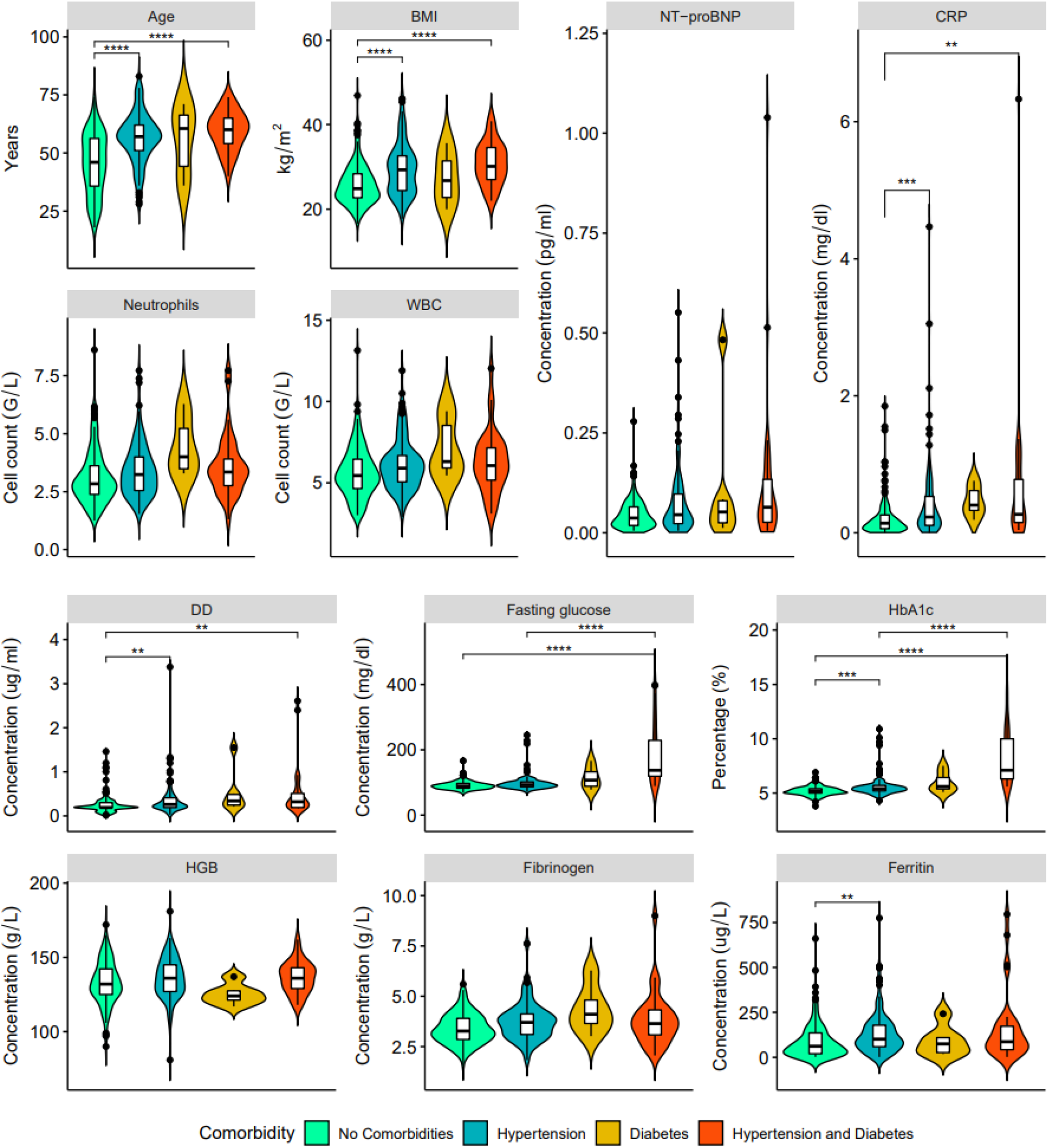
Clinical characteristics and laboratory results of blood samples in post-COVD-19 patients with no comorbidities (green), hypertension only (blue), type 2 diabetes mellitus only (orange) and both hypertension and type 2 diabetes (red): age, BMI, NT-proBNP, CRP, neutrophils, WBC, DD, fasting glucose, HbA1c, HGB, fibrinogen and ferritin (\*\**p* < 0.01, \*\*\**p* < 0.001, \*\*\*\**p* < 0.0001).

Multiple group comparisons had shown that from the parameters highlighted above, older age, higher body mass index, increased D-dimers, and NT-proBNP are primarily correlated with hypertension rather than diabetes mellitus. On the other hand, higher levels of fasting glucose, HbA1c, and C-reactive protein are attributed to having both comorbid states rather than one. Neutrophils, white blood cells, NT-pro BNP, hemoglobin, and fibrinogen were not significantly different among groups, according to the post-hoc test.

### 3.3 Lung Function Screening

#### Chest CT

At the follow-up visit with mean of 5.8±0.9 months chest CT was conducted in 227 subjects (72.8%). All the chest CT findings are reported in **Table 4** from the most common to the least common findings. 12 subjects (5.29%) had normal chest CT. The most common registered CT abnormality was chronic bronchitis in (198, 87.2%) followed by fibrotic changes in (83, 36.6%) and mediastinal lymphadenopathy in (23, 10.1%). Most of the participants had more than one CT changes. Median age of participants with lung abnormalities in chest CT was 55 years of age (IQR, 7). on 227 (72.8%) participants. Other CT abnormalities included polysegmental pneumonia, bronchiectasis, emphysema, bullous changes, atelectasis, and interstitial pneumonia (**Appendix 3**). These chest CT findings do not have a comorbidity-associate predisposition. However, fibrotic changes were found in 55 out of 158 patients with hypertension in comparison to 30 out of 154 participants without hypertension. (**Figure 5A**).

**Figure 5.**
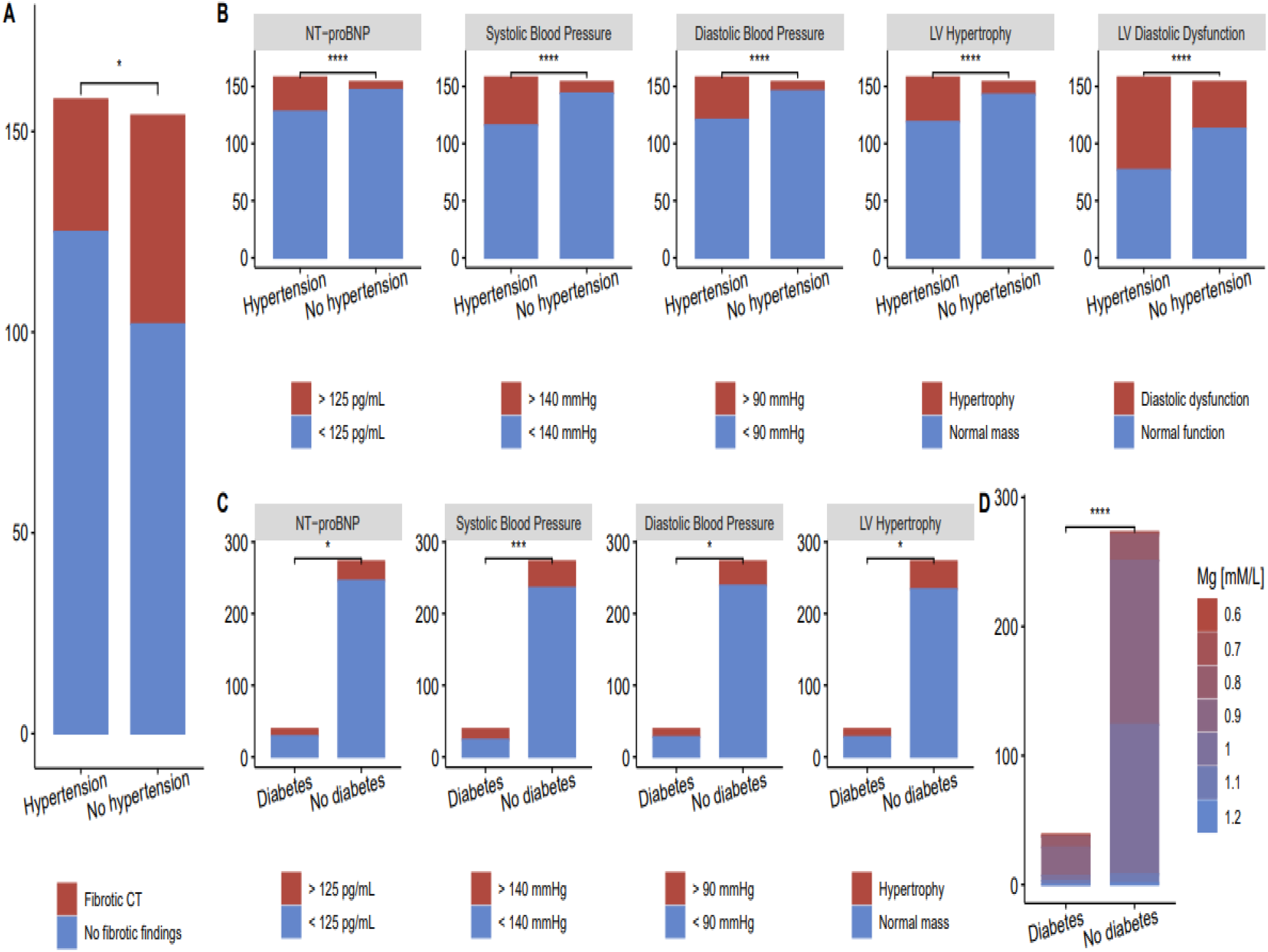
Fibrotic changes in CT scan of post-COVID-19 patients with hypertension (**A**). Cardiac abnormalities in post-COVID-19 patients with and without hypertension (**B**); with and without type 2 diabetes (**C**). (\**p* < 0.05, \*\*\**p* < 0.001, \*\*\*\**p* < 0.0001).

Overall, 107 participants had chest CT during active COVID-19 infection, with the mean percentage of lung defects 36.5±22.6%. Among those participants, 87 subjects (39.6%) underwent chest CT at the follow-up, and 84 (96.6%) subjects had residual lung abnormalities detected at the follow-up.

#### Spirometry test

All participants had spirometry test at the follow-up visit and 143 subjects (45.8%) had spirometry without abnormalities. From 169 subjects (54.2%) with abnormal spirometry 118 subjects (37.8%) had restrictive pattern and 43 subjects (13.8%) had mixed results with mostly restrictive pattern. Only 8 subjects (2.6%) had obstructive defects detected in spirometry. Regarding participants with respiratory comorbidities (9), there were 1 participant with restrictive, 1 participant with obstructive, and 4 participants with mixed with mostly restrictive defects. 67 participants (60.3%) out of 111 hospitalized participants had abnormal spirometry results.

#### Correlation between chest radiographs and 6MWT with lung function

125 (58.1%) out of 215 subjects with abnormal chest CT at the follow-up had abnormalities in spirometry test. 89 subjects (71.2%) had restrictive pattern, 30 subjects (24%) had mixed with mostly restrictive pattern, while 6 subjects (4.8%) had obstructive pattern in spirometry.

The median 6MWT in participants with abnormal spirometry was 381 (310;416) meters, significantly different from participants with normal spirometry (400 (350;450) meters, *P* = 0.004). 14 subjects (77.8%) out of 18 subjects with specific respiratory symptoms, including wheezing, dry cough, and cough with sputum, had abnormalities in chest CT at the follow-up. Meanwhile, 32 (68.9%) out of 45 subjects with dyspnea on exertion and 15 (83.3%) out of 18 subjects with dyspnea on rest had abnormal chest CT scan.

### 3.4 Cardiac screening

All 312 participants underwent a cardiac screening (**Table 3**). Sinus bradycardia <50bpm was present in 4 participants (1.28%). 37 participants (11.9%) had higher NT-pro BNP levels (> 125pg/ml) and left ventricular ejection fraction was decreased in 50 participants (1.60%). The most common cardiac abnormality was decreased diastolic function of the left ventricle (122, 39.1%). Left ventricular hypertrophy was present in all those participants. 50 (15.5%) had left ventricular hypertrophy, and 40 of them had preexisting hypertension. Meanwhile, 9 participants (18%) out of 50 with left ventricular hypertrophy did not have any cardiovascular comorbidities before the follow-up. Seven participants of those had left ventricular hypertrophy and diastolic dysfunction of LV, while 2 participants had dilatation of the left atrium in addition to left ventricular hypertrophy and diastolic dysfunction.

**Table 3.** Cardiac abnormalities detected in cardiac assessment of post-COVID-19 patients.

|  |  |  |
| --- | --- | --- |
| **Serum cardiac biomarker** | Total |  |
| NT-proBNP > 125pg/ml | 37 (11.9) |  |
| **Electrocardiography** |  |  |
| Sinus bradycardia <60bpm | 26 (8.33) |  |
| Sinus bradycardia <50bpm | 4 (1.28) |  |
|  | Total | Newly diagnosed |
| **Echocardiography** |  |  |
| Left ventricular ejection fraction<sup>†</sup>, % | 61±4.55 |  |
| Left ventricular ejection fraction <50% | 5 (1.60) |  |
| Left ventricular hypertrophy | 50 (15.5) | 7 (2.24) |
| Diastolic dysfunction of left ventricle | 122 (39.1) | 2 (0.64) |
| Dilation of right atrium | 2 (0.64) | 1 (0.32) |
| Dilation of left atrium | 12 (3.85) |  |
| **Holter electrocardiography** |  |  |
| Atrial Fibrillation | 5 (1.60) | 4 (1.28) |
| AV block II degree | 4 (0.32) | 3 (0.96) |
| Sinoatrial block | 1 (0.32) | 1 (0.32) |
| RBBB | 1 (0.32) | 1 (0.32) |
| **24-hr BPM** |  |  |
| Mean SBP ≥140mmHg | 71 (22.8) | 19 (6.10) |
| Mean DBP ≥90 mm Hg | 49 (15.7) | 19 (6.10) |
| Note. – Except where indicated, data are numbers of participants, with percentages in parentheses. |  |  |
| 24-hr BPM – 24-hour blood pressure monitoring |  |  |
| † data are means ± standard deviation |  |  |

Holter ECG revealed heart rhythm disturbances in 9 participants for the first time (2.88%), including atrial fibrillation (4, 1.28%), AB block II degree (3, 0.96%), SA block (1, 0.32%), and RBBB (1, 0.32%). The median age of patients with the first onset of rhythm disturbances was 69 years (IQR, 12), and no difference in gender was found. 2 patients with new-onset AF had preexisting hypertension, and one patient had newly reported hypertension at the follow-up.

The mean systolic blood pressure (SBP) of all participants was 120.7±16. mm Hg, and the mean diastolic blood pressure (DBP) was 78.4±9.03mmHg with no significant difference between females and males. Participants with elevated sBP defined as ≥140 mmHg were present in 71 subjects (22.8%).

24-hour blood pressure monitoring revealed 19 patients (6.10%) with elevated blood pressure either in the daytime or nighttime for the first time. The median age of participants with new-onset hypertension was 45 years (IQR, 12). One participant was pregnant during the screening, and none of the patients had cardiovascular conditions before the follow-up time. Two patients had type 2 diabetes. One patient had LV hypertrophy, 7 had diastolic dysfunction of LV, one patient had dilation of RA, and one patient was with dilation with LA.

Cardiac abnormalities in patients with hypertension and diabetes type 2 compared to participants with no comorbidities are shown in **Figure 5B** and **5C** respectively. 24-hour blood pressure monitoring revealed significantly higher systolic and diastolic blood pressure, left ventricular hypertrophy and elevated NT-proBNP in participants with hypertension and subjects with type 2 diabetes. Left ventricular diastolic dysfunction more frequently present in patients with hypertension.

#### Laboratory results

The results of blood laboratory analysis at the follow-up are reported in **Table 4** and **Figures 2-4**. The minority of subjects had persistent inflammatory markers at the follow-up, including neutrophilia in 18 subjects (5.77%) and increased CRP levels in 54 subjects (17.3%). 28 subjects (8.97%) had leukopenia with WBC < 4×109/l and 13 subjects (4.17%) had lymphopenia with lymphocytes<1.18x 103/l.

**Table 4.**
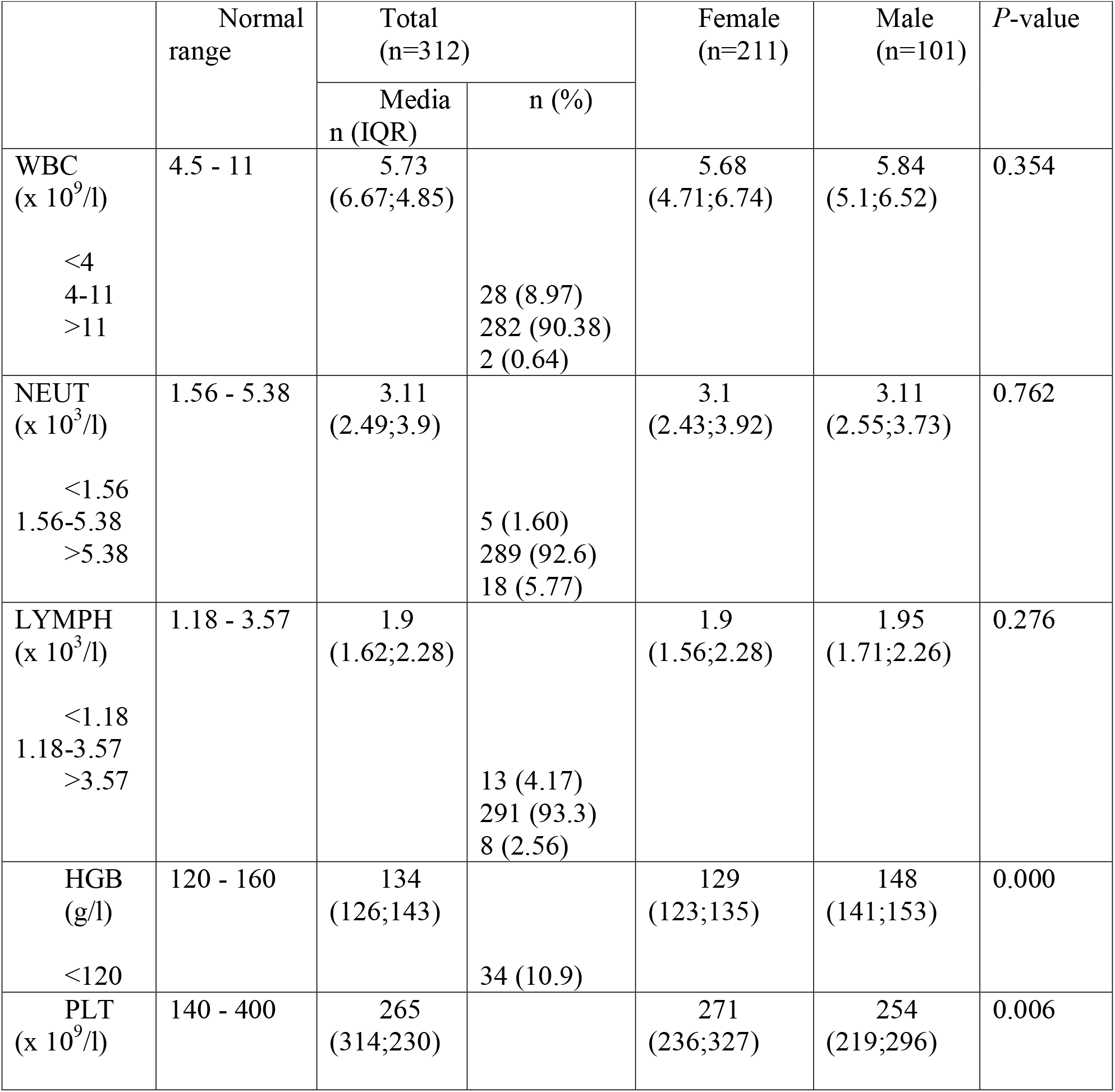
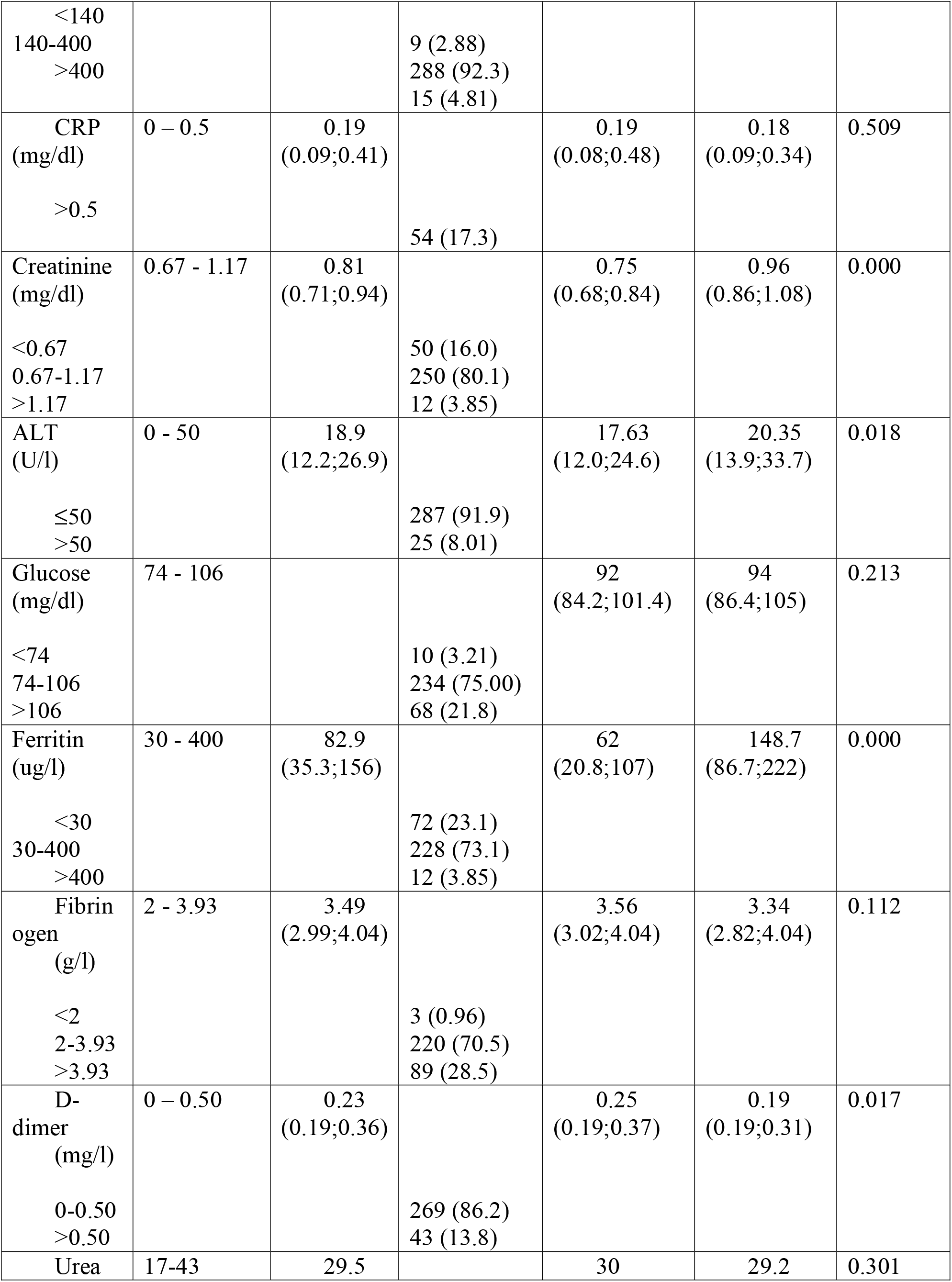
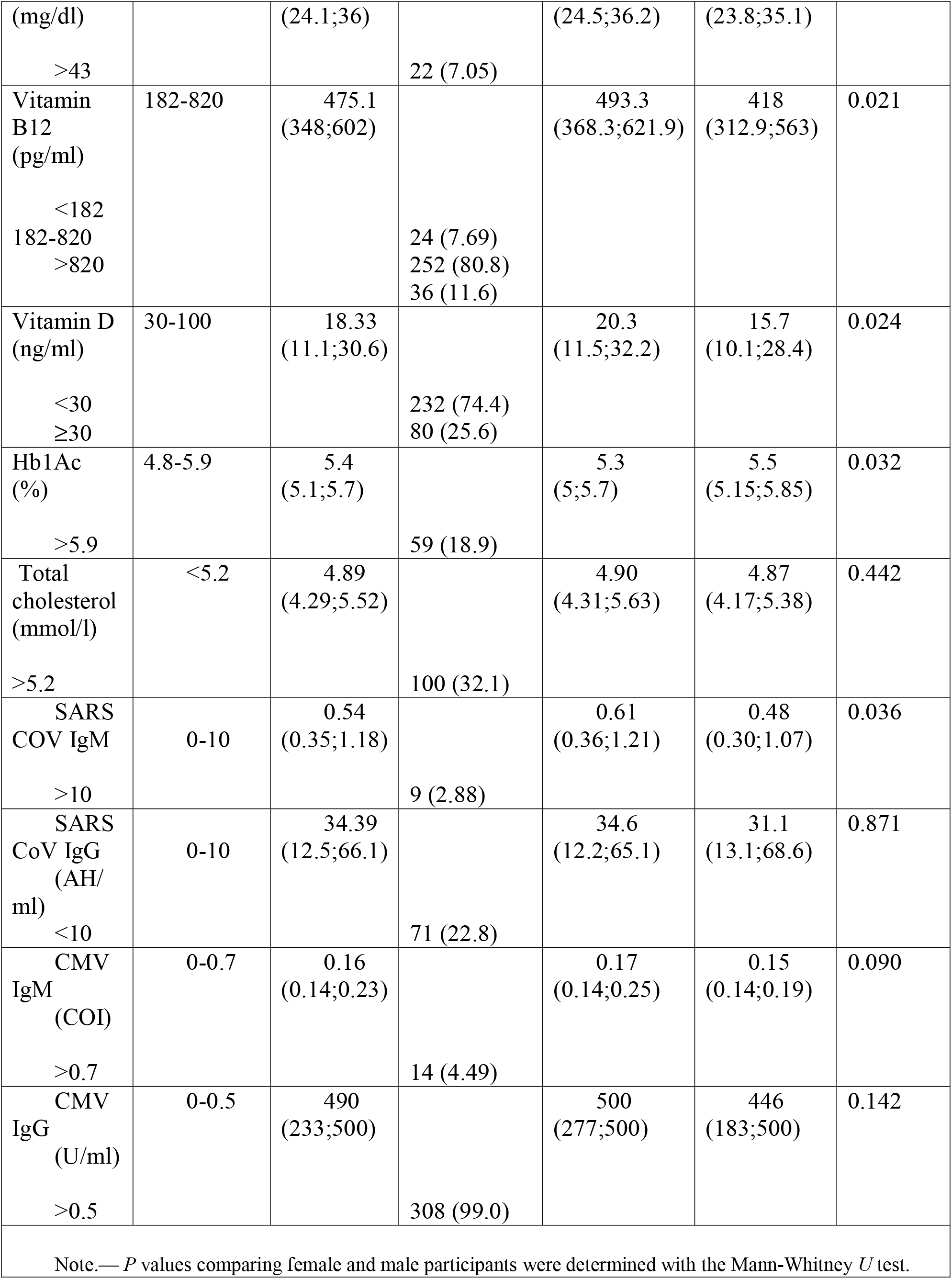
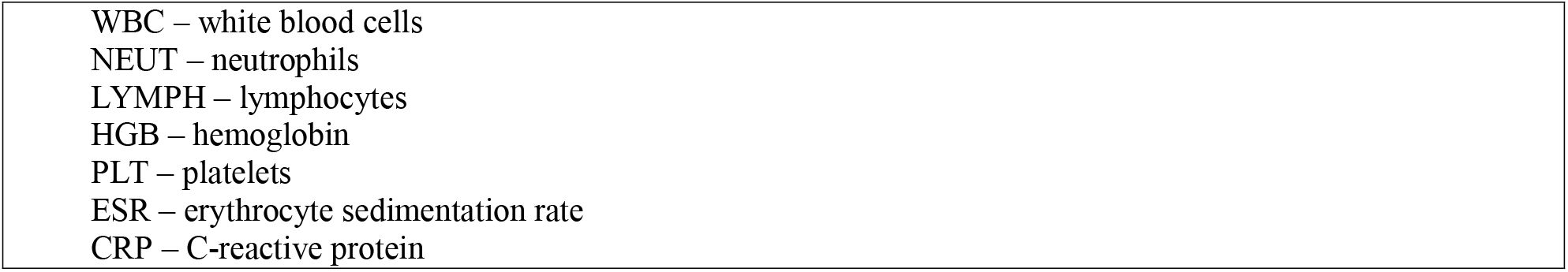
Laboratory results of blood sample in participants after COVID-19 infection with median follow-up of 6 months.

|  | Normal range | Total (n=312) |  | Female (n=211) | Male (n=101) | P-value |
| --- | --- | --- | --- | --- | --- | --- |
|  |  | Media n (IQR) | n (%) |  |  |  |
| WBC (x 10<sup>9</sup>/l) | 4.5 - 11 | 5.73 (6.67;4.85) |  | 5.68 (4.71;6.74) | 5.84 (5.1;6.52) | 0.354 |
| <4 |  |  | 28 (8.97) |  |  |  |
| 4-11 |  |  | 282 (90.38) |  |  |  |
| >11 |  |  | 2 (0.64) |  |  |  |
| NEUT (x 10<sup>3</sup>/l) | 1.56 - 5.38 | 3.11 (2.49;3.9) |  | 3.1 (2.43;3.92) | 3.11 (2.55;3.73) | 0.762 |
| <1.56 |  |  | 5 (1.60) |  |  |  |
| 1.56-5.38 |  |  | 289 (92.6) |  |  |  |
| >5.38 |  |  | 18 (5.77) |  |  |  |
| LYMPH (x 10<sup>3</sup>/l) | 1.18 - 3.57 | 1.9 (1.62;2.28) |  | 1.9 (1.56;2.28) | 1.95 (1.71;2.26) | 0.276 |
| <1.18 |  |  | 13 (4.17) |  |  |  |
| 1.18-3.57 |  |  | 291 (93.3) |  |  |  |
| >3.57 |  |  | 8 (2.56) |  |  |  |
| HGB (g/l) | 120 - 160 | 134 (126;143) |  | 129 (123;135) | 148 (141;153) | 0.000 |
| <120 |  |  | 34 (10.9) |  |  |  |
| PLT (x 10<sup>9</sup>/l) | 140 - 400 | 265 (314;230) |  | 271 (236;327) | 254 (219;296) | 0.006 |
| <140<br>140-400<br>>400 |  |  | 9 (2.88)<br>288 (92.3)<br>15 (4.81) |  |  |  |
| CRP<br>(mg/dl)<br>>0.5 | 0 – 0.5 | 0.19<br>(0.09;0.41) | 54 (17.3) | 0.19<br>(0.08;0.48) | 0.18<br>(0.09;0.34) | 0.509 |
| Creatinine<br>(mg/dl)<br><0.67<br>0.67-1.17<br>>1.17 | 0.67 - 1.17 | 0.81<br>(0.71;0.94) | 50 (16.0)<br>250 (80.1)<br>12 (3.85) | 0.75<br>(0.68;0.84) | 0.96<br>(0.86;1.08) | 0.000 |
| ALT<br>(U/l)<br>≤50<br>>50 | 0 - 50 | 18.9<br>(12.2;26.9) | 287 (91.9)<br>25 (8.01) | 17.63<br>(12.0;24.6) | 20.35<br>(13.9;33.7) | 0.018 |
| Glucose<br>(mg/dl)<br><74<br>74-106<br>>106 | 74 - 106 |  | 10 (3.21)<br>234 (75.00)<br>68 (21.8) | 92<br>(84.2;101.4) | 94<br>(86.4;105) | 0.213 |
| Ferritin<br>(ug/l)<br><30<br>30-400<br>>400 | 30 - 400 | 82.9<br>(35.3;156) | 72 (23.1)<br>228 (73.1)<br>12 (3.85) | 62<br>(20.8;107) | 148.7<br>(86.7;222) | 0.000 |
| Fibrin<br>ogen<br>(g/l)<br><2<br>2-3.93<br>>3.93 | 2 - 3.93 | 3.49<br>(2.99;4.04) | 3 (0.96)<br>220 (70.5)<br>89 (28.5) | 3.56<br>(3.02;4.04) | 3.34<br>(2.82;4.04) | 0.112 |
| D-dimer<br>(mg/l)<br>0-0.50<br>>0.50 | 0 – 0.50 | 0.23<br>(0.19;0.36) | 269 (86.2)<br>43 (13.8) | 0.25<br>(0.19;0.37) | 0.19<br>(0.19;0.31) | 0.017 |
| Urea | 17-43 | 29.5 |  | 30 | 29.2 | 0.301 |
| (mg/dl) |  | (24.1;36) |  | (24.5;36.2) | (23.8;35.1) |  |
| >43 |  |  | 22 (7.05) |  |  |  |
| Vitamin B12 (pg/ml) | 182-820 | 475.1 (348;602) |  | 493.3 (368.3;621.9) | 418 (312.9;563) | 0.021 |
| <182 |  |  | 24 (7.69) |  |  |  |
| 182-820 |  |  | 252 (80.8) |  |  |  |
| >820 |  |  | 36 (11.6) |  |  |  |
| Vitamin D (ng/ml) | 30-100 | 18.33 (11.1;30.6) |  | 20.3 (11.5;32.2) | 15.7 (10.1;28.4) | 0.024 |
| <30 |  |  | 232 (74.4) |  |  |  |
| ≥30 |  |  | 80 (25.6) |  |  |  |
| Hb1Ac (%) | 4.8-5.9 | 5.4 (5.1;5.7) |  | 5.3 (5;5.7) | 5.5 (5.15;5.85) | 0.032 |
| >5.9 |  |  | 59 (18.9) |  |  |  |
| Total cholesterol (mmol/l) | <5.2 | 4.89 (4.29;5.52) |  | 4.90 (4.31;5.63) | 4.87 (4.17;5.38) | 0.442 |
| >5.2 |  |  | 100 (32.1) |  |  |  |
| SARS COV IgM | 0-10 | 0.54 (0.35;1.18) |  | 0.61 (0.36;1.21) | 0.48 (0.30;1.07) | 0.036 |
| >10 |  |  | 9 (2.88) |  |  |  |
| SARS CoV IgG (AH/ml) | 0-10 | 34.39 (12.5;66.1) |  | 34.6 (12.2;65.1) | 31.1 (13.1;68.6) | 0.871 |
| <10 |  |  | 71 (22.8) |  |  |  |
| CMV IgM (COI) | 0-0.7 | 0.16 (0.14;0.23) |  | 0.17 (0.14;0.25) | 0.15 (0.14;0.19) | 0.090 |
| >0.7 |  |  | 14 (4.49) |  |  |  |
| CMV IgG (U/ml) | 0-0.5 | 490 (233;500) |  | 500 (277;500) | 446 (183;500) | 0.142 |
| >0.5 |  |  | 308 (99.0) |  |  |  |
| Note.— *P* values comparing female and male participants were determined with the Mann-Whitney *U* test. |  |  |  |  |  |  |

|  |
| --- |
| WBC – white blood cells |
| NEUT – neutrophils |
| LYMPH – lymphocytes |
| HGB – hemoglobin |
| PLT – platelets |
| ESR – erythrocyte sedimentation rate |
| CRP – C-reactive protein |

Immunological test results showed that SARS-CoV19 IgM antibodies were present in 9 subjects (2.88%). Among those, 5 participants had follow-up at 5th months after the onset of the disease, while the other 4 participants had follow-up at 6th months. SARS-CoV19 IgG antibodies were present in 241 subjects (77.2%) and were <10 AH/ml in 71 participants (22.8%). Meanwhile, increased CMV IgG titers defined as >0.5 U/ml were present in almost all 312 participants.

## 4. Discussion

In our study 6 months after acute illness with COVID-19, most of patients still meet the criteria for long COVID-19 and show at least one persistent symptom. These results are mainly in line with studies reporting the prevalence of long COVID-19 in patients 3 to 6 months after COVID-19 (Al-Aly et al., 2021; Huang C et al., 2021; Nasserie et al., 2021; Nehme et al., 2021). Long-Covid-19 symptoms have similarities with chronic fatigue syndrome developed after viral infections (Holmes et al., 1988; Hickie et al., 2006) and may afflict a wide range of organs, with the most common symptoms being chronic fatigue, shortness of breath, cognitive impairments (Cares-Marambio et al., 2021; Shah et al., 2021; Davido et al., 2020; Halpin et al., 2020).

The pathophysiology of long COVID-19 respiratory and multi-organ sequelae has a fundamental knowledge gap that must be addressed (Lerner et al., 2021). Viral infections are considered to be cofactors for the initiation and exacerbation of lung fibrosis (Naik, Moore, 2010), and clinical evidence suggesting cardiovascular involvement has been reported for essentially all known viral infections (Abelmann, 1971). Observational studies revealed a high percentage of post-COVID-19 sequelae such as respiratory problems, lung fibrosis (Lechowicz et al., 2020; Ali et al., 2021), and cardiovascular problems (Bose, McCarthy, 2020). Moreover, long COVID-19 affects even mild-to-moderate cases of survivors of COVID-19 with pulmonary sequelae reported after recovery (Wang et al., 2020). Secondly, our study demonstrated that deranged blood indices (increased neutrophils, WBC, changed coagulation profile-significantly increased DD; fasting glucose, HbA1c, HGB) and raised inflammatory marker (CRP) are associated with pre-existing comorbidities, specifically, hypertension and diabetes. HbA1c is considered the gold standard for evaluating blood glucose levels (Nalysnyk et al., 2010) and is considered to be associated with inflammation, hypercoagulability, blood oxygen saturation, and higher mortality rate in COVID-19 patients (Wang et al., 2020).

Hypertension affects more than a quarter of the global population (Kearney et al., 2005; Forouzanfar et al., 2017), has been identified as a major risk factor for COVID-19 severity (Liang et al., 2020; Madhur et al., 2021) and higher risk of dying from COVID-19 (Gao et al., 2020). Moreover, hypertension is closely associated with other comorbidities, predominantly obesity and diabetes, and significantly affects inflammation (Jayedi et al., 2019). In this study, hypertension, besides the above-mentioned significant changes in blood and inflammatory markers that coincided with other comorbidities (diabetes), also revealed significant changes in fibrinogen (Fbg) and ferritin levels. Changes in Fbg levels can be associated with a substantial rearrangement in blood vessel walls observed in individuals with pre-existing hypertension. The abnormal Fbg levels have a strong association to COVID-19 disease severity (rev. Len et al., 2022), and assessment of Fbg levels at admission is important for determining of prognosis of COVID-19 patients (Long et al., 2021). Abnormal Fbg levels are observed in post-COVID-19 cohorts by different groups (Tartari et al., 2021). Cardiovascular changes associated with COVID-19 may lead to left ventricular dysfunction (Medranda et al., 2021), which in our cohort had a higher prevalence in convalescent patients with hypertension.

Cardiac screening showed a mean ejection fraction 61±4.55%, which is less than in participants observed 1-4 weeks after discharge (Zhou, 2021). This can be explained by a higher prevalence of chronic heart diseases, including hypertension and CAD, in our cohort of participants. Interestingly, our study revealed new cases of left ventricular hypertrophy and diastolic dysfunction of LV in participants with no preexisting cardiovascular disorders. In addition, 19 participants had newly diagnosed hypertension during the follow-up. The patients with newly diagnosed hypertension were young (median, 45 years) and had no preexisting cardiovascular comorbidities before the follow-up. However, echocardiography revealed long-term complications in some of those patients, meaning that hypertension might have been before the 6-month follow-up and COVID-19 infection. In addition to hypertension and LV dysfunction, our study found 9 new cases of rhythm disturbances detected in 24-hour BP monitoring. Newly detected rhythm disturbances included atrial fibrillation (AF), AB block II degree, sinoatrial (SA) block, and RBBB (**Table 3**).

Although our cohort was limited to establishing the time of the onset of hypertension in the post-COVID-19 period, our findings in patients with no preexisting cardiac comorbidities and standard echocardiography suggest the presence of the long-term effect of COVID-19 on cardiac function. The evidence of cardiac injury in post-COVID-19 patients (left ventricular ejection fraction and diastolic dysfunction) and the appearance of arrhythmias corroborate with the findings of other investigators (rev. Satterfield et al., 2022; Tudoran et al., 2022). The arrhythmias in post-COVID patients were found to be associated with COVID-19 disease severity (Mohammad et al., 2021).

In patients with COVID-19, the viral infection may initiate inflammation with subsequent fibrosis, and cardiac inflammation and fibrosis are major pathological mechanisms leading to heart failure (Suthahar et al., 2017). Non-invasive imaging, blood analysis for fibrosis markers, and endomyocardial biopsy (EMB) are used to detect fibrosis (Raafs et al., 2021). The primary imaging technique in contemporary heart failure management is echocardiography; however, it provides little information about the extent of fibrosis (de Boer et al., 2019). To evaluate the cardiac and lung changes we used echocardiography and the chest CT.

The question if the lung is a target organ for diabetes mellitus has been discussed, and a body of clinical and experimental evidence indicates that fibrosis and other pulmonary complications are more prevalent than it was early recognized (Sandler, 1990; Hsia, Raskin, 2005; Yang et al., 2011). Critically fibrotic changes on follow-up CT scans were significantly associated with hypertension but not diabetes (**Figure 5**). The presence of diabetes and new-onset diabetes was an independent factor associated with poor outcomes during different coronavirus infections – SARS-CoV-1 (Yang et al., 2006), MERS-CoV (Alraddadi et al., 2016), and SARS-CoV-2 (rev. Singh, Singh, 2020). In our study, hypertension had a significant association with fibrotic changes on CT-scan, whereas diabetes type II did not. One of the possible explanations can be a small percentage of patients with diabetes type II in our Long COVID-19 cohort. However, we may also speculate that the fibrotic changes in post-COVID-19 develop faster when blood vessels are already affected by hypertension. It was reported that abnormal lung CT findings could present even in asymptomatic or mild COVID-19 patients (Shi et al., 2020). The association of radiological CT findings with hypertension in Long COVID-19 makes hypertension, in combination with other results (high fibrinogen, DD, etc.), a possible early marker of developing fibrotic lung changes.

Laboratory tests identified the long-term persistence of IgM antibodies in 9 patients from our cohort (5-6 months). The IgM antibodies at COVID-19 infection peak early and rapidly decline (rev. Huang AT et al., 2020; Mallon et al., 2021; Štěpánek et al., 2022). IgM seropositivity was found to be correlating with a symptom duration (Štěpánek et al., 2022). Moreover, the long-term persistence of anti-SARS-CoV-2 IgM antibodies was recently reported by Bichara and colleagues (2021), who observed a long 8-months persistence of the IgM antibodies after COVID-19 infection in two patients. The prolonged persistence of anti-viral IgM antibodies was described after different acute viral infections and live viral vaccines: the Japanese encephalitis live vaccine (Hills et al., 2021), acute hepatitis A infection (Kao et al., 1984), dengue virus infection (Chien et al., 2018), Zika virus > 2 years (Griffin et al. 2019), yellow fever virus vaccine > 3-4 years (Gibney et al., 2012), and in cerebrospinal liquid and blood of patients after West Nile virus infection (Kapoora et al., 2004; Roehrig et al., 2003; Papa et al., 2011; Murray et al., 2013) in some cases up to 8 years after infection. The finding of long persistence of IgM antibodies in some participants of the post-COVID-19 cohort warrants additional investigation, particularly the determination of whether persistence of IgM is related to persistent infection with SARS-CoV2. However, the finding of elevated IgG anti-CMV antibodies in most of the cohort should be interpreted with caution due to the possibility of cross-reactivity between viral epitopes and human tissue antigens (Kreye et al., 2020; Vojdani et al., 2021).

According to Chalder Fatigue Score, the prevalence of some symptoms, as well as neurological symptoms had a significant association with the female gender. This finding corroborates the recent conclusions of another prospective study of Long COVID-19 syndrome (Bai et al., 2022).

There are several limitations of our study: (1) Patients only with persistent symptoms were selected for the study, and there was no control group for comparison of patients with and without the Long COVID-19 syndrome; (2) The absence of baseline health data and CT-imaging data before the infection and before the mean of 6 months of follow-up limits the assessment of changes in patient’s results; (3) Social information of patients, including the number of years of smoking, alcohol use, and lifestyle, was not reported, which might have a confounding effect on some of the results. We did not include in the asymptomatic cohort individuals with a mild phenotype of disease though non-hospitalized subjects with moderate post-COVID-19 were included; (4) We had some limitations on cardiovascular measures. It would not be appropriate or feasible to obtain histological data from otherwise healthy subjects. The data in this study permit a 6-months assessment of Long-COVID-19 sequelae. Long-term prospective studies of longer duration are still needed.

## 5. Conclusions

In the 6-months follow up Long COVID-19 cohort, among the tested parameters, those positively correlated with comorbid conditions include age, BMI, D-dimers, NT-proBNP, C-reactive protein, neutrophils, fasting glucose, and HbA1c; hypertension also shows three associations that were not found in patients when examining the role of diabetes: increased hemoglobin, fibrinogen, and ferritin. In addition to hypertension and LV dysfunction, our study found new cases of rhythm disturbances detected in 24-hour BP monitoring that included atrial fibrillation (AF), AB block II degree, sinoatrial (SA) block, and RBBB. We speculate that due to the significant association of hypertension with CT fibrotic changes, it may serve as an early predictive marker (in combination with high Fbg and DD) for the development of lung parenchymal diseases in post-COVID-19.

With the new outbreaks of SARS-CoV-2 infection in many countries, it is expected that the patients’ burden having Long COVID-19 respiratory and cardiovascular sequelae is going to increase and lead to another public health crisis following the current pandemic. Thus, it is vital for further research comprising longer-ranging observational studies critical to evaluate the long-term consequences of COVID-19. The preliminary study presented here can be prolonged to 12 and 24 follow-ups. This will enable us to study the long-term effects of Long COVID-19 and identify symptoms significant for medical treatment and observation.

## Data Availability

All data produced in the present study are available upon reasonable request to the authors

## Acknowledgements

This research has been funded by the Science Committee of the Ministry of Education and Science of the Republic of Kazakhstan (Grant No. BR10965164). https://www.ncste.kz/en/main. The funders had no role in study design, data collection and analysis, decision to publish, or preparation of the manuscript.

## Authors’ contributions

AT, ZK, BA, NK, MB - Conceptualization, Investigation, Methodology, Supervision, Writing – review & editing, AS, SK - Data curation, project administration. ZS, PL – Methodology, Statistical analysis. NSB – Supervision, Analysis, Writing a draft. All authors read and edit a final draft. All authors contributed to the article and approved the submitted version.

## Data Availability Statement

There is a risk of identification of sensitive data of individual patients due to small subgroups; therefore, the data are not openly accessible on request.

## Abbreviations

BMI: body mass index
CAD: coronary artery disease
COPD: chronic obstructive pulmonary disease
CRP: C-reactive protein
CMV: cytomegalovirus
CT: computer tomography
DBP: low diastolic blood pressure
DD: D-dimer
Fbg: fibrinogen
GI disorders: gastrointestinal disorders
HbA1c: glycohemoglobin A1c
LV: Left ventricular
6MWT: six minute test
NT-proBNP: ventricular natriuretic peptide
RBBB: right bundle branch block
SA: sinoatrial
SBP: systolic blood pressure

## Supplemental Data

**Appendix 1**. Results of a questionnaire with items on demographic information, current complaints, comorbidities, and medications.

**Appendix 2**. Results of a Chalder Fatigue Scale (CFS) questionnaire.

**Appendix 3**. CT abnormalities.

